# Implementation of a standardized Video-based Asynchronous Neurological Examination (VANE) in a multi-center observational study of Alzheimer’s disease (AD) and AD related dementias

**DOI:** 10.64898/2026.07.15.26357456

**Authors:** JM Noble, NK Nadkarni, D Martinez, M Temprosa, A Bowers, O Carmichael, L Doherty, GJ Febres, D Sanchez, TE Goldberg, H Sherif, Vallabh Shah, JA Luchsinger, the Diabetes Prevention Program Research Group

**Author notes:** Corresponding author: James M. Noble MD, MS, CPH, FAAN, Professor of Neurology at CUMC, Department of Neurology, Taub Institute for Research on Alzheimer’s Disease and the Aging Brain, and the GH Sergievsky Center, Columbia University, c/o Diabetes Prevention Program Coordinating Center George Washington University Biostatistics Center 6710A Rockledge Drive, Suite 250, Bethesda Maryland 20817, Tele, www.dppos.org. Acknowledgement: The DPP Research Group gratefully acknowledges the commitment and dedication of the participants of the DPP and DPPOS.

## Abstract

**Introduction:** The Diabetes Prevention Program Outcomes Study (DPPOS) is an established cohort of aging persons with pre-diabetes and type 2 diabetes with 25 years of median follow-up. In 2022 DPPOS added Alzheimer’s disease (AD), and AD related dementias (ADRD) phenotyping using the National Alzheimer’s Coordinating Center (NACC) Uniform Data Set (UDSv3), which included a standardized neurological examination across 25 clinical sites, administered by clinical staff and interpreted centrally by clinicians.

**Methods:** A DPPOS video-based asynchronous neurological examination (DPPOS-VANE) was developed iteratively through consensus from research clinicians and staff feedback to harmonize with UDSv3 to identify common neurological diagnoses aside from dementia including diabetic cranial neuropathies, stroke and parkinsonism. DPPOS-VANE was designed to be conducted without direct participant contact by the examiner, reproducible, and independent of clinical skills of PCs. An iPad™ camera recorded the video exam, comprised of assessments of extraocular and facial movements, visual fields, speech, gross motor strength, pronator drift, praxis and parkinsonism. A 10-minute training video demonstrated the examination step-by-step with scripts and instructions in English and Spanish. Site-specific performance review, feedback, and staff certification preceded central reading of video recordings by physicians.

After two years of implementation, 1286 DPPOS-VANEs led to 1284 examination reviews. Of these, 1204 (93%) were completed by having the examiner follow the standard script. Overall, 1237 examinations (96%) were delivered as planned, 41 (3%) had minor errors but were still usable, and 6 (0.4%) had major deviations in exam technique; two additional recorded evaluations were not usable as recorded videos were inaccessible due to technical errors. Each examination was completed within 10-15 minutes. Each site on average completed 51.4 examinations (range 14-92).

**Discussion:** Engaging 55 research staff across 25 sites and 3 physician-reviewers, this study is the first to demonstrate feasibility of a VANE as an efficient neurological examination model enabled by commonly used devices. Such a multisite standardized VANE represents a novel paradigm for large epidemiological studies.

## Introduction

There is an urgent need to expand knowledge of how neurological diseases manifest with age across different populations, particularly as they may reflect secular trends of treatments of risk factors for neurological disease.^1^ This will require leveraging large multisite studies. The largest national epidemiological studies are powered to detect small differences across multiple population segments, whether or not they are combined with brief questionnaires and biological phenotyping on a small segment for imputation to others.^2,3^

Historically, frameworks of basic, screening, or comprehensive neurological examinations^4,5^ have been included in large national studies of aging populations only when cognition was a major outcome or focus of the study itself.^6–8^ In contrast, focused neurological examinations may be included as an element of a study when a particular problem is of interest to the investigators, such as peripheral neuropathy,^9^ particularly in populations with or at high risk for diabetes mellitus.^10^ Moreover, neurological examinations are burdensome, adding at least 5 minutes of time to clinical evaluations among practicing neurologists, underscoring the challenge of implementing efficient neurological assessments even in settings with experienced evaluators.^11^

Neurological examinations in cognitive aging studies traditionally take place through in-person assessments conducted by expert trained clinicians.^6–8,12^ This requires substantial clinical personnel including a physician or advance practice provider (APP, including physician associates and nurse practitioners) with sufficient examination expertise. In contrast, neuropsychological testing is commonly delivered by research assistants (RA) and project coordinators (PC) who have been trained by local neuropsychologists. However, neurological examinations conducted and recorded by RA and PC have not traditionally been considered feasible.

In studies conducting neuropsychological testing leading to cognitive adjudications and diagnoses, combining these with neurological examination findings becomes increasingly important in assessing a clinic-biological diagnostic framework of Alzheimer’s Disease (AD) and AD-related dementias (ADRD). Neurological problems including stroke and parkinsonism can be subtle and are often missed on examination, and become increasingly common with advancing age.^13^ Parkinsonism, apraxia, gaze palsies and gait abnormalities are elements of parkinsonian disorders, stroke, vascular dementia and other ADRD. Inclusion of such findings from neurological examinations is necessary for precise cognitive outcome adjudications. The importance of the neurological examination is among the reasons that the National Alzheimer’s Association Coordinating Center (NACC) Unified Data Set (NACC-UDS) incorporates neurological clinical assessments, aiming to document neurologic findings most relevant to cognitive diagnoses.

With the advent of remote video care, accelerated during the COVID-19 pandemic, new models have emerged to enable remote neurological examinations.^2,14^ Here we present the development and implementation of a video-based asynchronous neurological examination (VANE) implemented in 25 nationwide sites of The Diabetes Prevention Program Outcomes Study Phase 4 (DPPOS-4), focused on AD/ADRD. VANE complemented other DPPOS-4 procedures including neuropsychological testing and multimodal biomarker-based assessments. VANE enabled completion of the neurological exam forms of the NACC-UDS version 3 forms.^6^ These forms require classifying persons as having (a) a normal neurological exam or (b) findings accompanying AD/ADRD or (c) findings not related to AD/ADRD. Findings consistent with AD/ADRD emphasize signs of cerebrovascular disease, parkinsonian syndromes, and other neurodegenerative diseases.

## Methods

### Study setting

The eligibility criteria, design, and methods of the Diabetes Prevention Program (DPP) ^15^ and Diabetes Prevention Program Outcomes Study (DPPOS)^16^ have been reported elsewhere. DPP was a randomized trial comparing the effects of an intensive lifestyle intervention (ILS), metformin, and placebo on diabetes incidence among 3,234 participants enrolled between 1996 and 1999. Participants were required to have a BMI ≥ 24 kg/m² (≥22 kg/m² in Asian Americans), fasting plasma glucose levels between 95 and 125 mg/dl, and impaired glucose tolerance (2-hour post-load glucose of 140–199 mg/dl). People were excluded if taking medications known to alter glycemia or if they had illnesses that could reduce their life expectancy or their ability to participate in the trial. Written informed consent was obtained from all participants before screening, consistent with the Declaration of Helsinki and the guidelines of each center’s institutional review board.

Beginning in year 8 of DPPOS (2009-2010), brief neuropsychological assessments were first introduced as outcomes and these have been tracked longitudinally.^17^ DPPOS Phase 4 was funded in 2022 with an emphasis on AD/ADRD phenotyping in the DPPOS cohort of persons with pre-diabetes and T2D, which required the implementation of the NACC-UDS version 3 forms, including the neurologic exam.

### VANE development

A video-based asynchronous neurological examination (VANE) was developed iteratively through consensus of AD/ADRD clinicians and feedback from DPPOS PCs to harmonize with the NACC-UDS Version 3 (UDSv3) Neurological Examination Findings (B8),^6^ which captures common neurological diagnoses (aside from AD/ADRD) including cranial neuropathies, stroke and parkinsonism.^13,18^ UDSv3 evaluations are expected to be conducted to address all B8 form questions, and by a clinician with experience in “assessing the neurological signs…and in attributing the observed findings to a particular syndrome.”^12^ The B8 form begins with a broad determination of a) no abnormal findings; b) abnormal findings associated with neurological conditions including parkinsonism, stroke, higher cortical dysfunction to suggest posterior cortical atrophy or apraxia of gaze, progressive supranuclear palsy or related atypical parkinsonians syndromes (e.g. corticobasal syndrome), amyotrophic lateral sclerosis, normal pressure hydrocephalus, or other neurological findings such as cerebellar ataxia, chorea, or myoclonus; or c) abnormal findings seen in aging but not traditionally associated with dementia (e.g., Bell’s palsy). As a means to save time, the VANE can be implemented in the delayed recall period of cognitive testing during which no cognitive task should be administered. In DPPOS-4, research staff were encouraged to conduct the delayed recall period of the Spanish-English Verbal Learning Test (SEVLT)^19^ as part of DPPOS-4 procedures, although participants could be tested at other times. The VANE also took advantage of the availability of iPADs for the administration of the NIH toolbox. iPads have video recording capability in addition to internet connectivity for direct video uploads.

General principles of the DPPOS-4 VANE design included no contact between the administrator and the participant, reproducibility, and reliability for interpretation, independence of clinical skills of DPPOS PCs, and no redundancy with other DPPOS-4 procedures. PC qualifications ranged from bachelor’s degrees to nursing degrees and doctoral degrees. All VANE acquisition relied on study site research staff for in-person examinations, followed by asynchronous interpretation by clinicians supported by the DDPOS-4 Cognitive Assessment and Adjudication Core at Columbia University Irving Medical Center. A dedicated core manager supported the clinicians and was the liaison with the DPPOS-4 coordinating center at George Washington University (CC-GWU) and the DPPOS-4 sites. VANE findings could be shared back with DPPOS-4 sites for quality control and reporting neurological abnormalities requiring attention.

Ethical safeguards were central to the VANE design and workflow. The asynchronous and centralized nature of video review creates a separation between examination and evaluation that is unique to this approach, and requires that consent procedures, data security, and accountability structures be explicitly defined at the outset of any implementation. In DPPOS, these were addressed through IRB oversight, restricted platform access, and a clearly defined chain of responsibility for the return of findings to participants. Prior to implementation for clinics, extensive multi-phase pilot testing was conducted to ensure a smooth and successful workflow in 3 dimensions: (1) technical feasibility and latency to ensure high quality of videos and MIDAS platform stability; (2) clinical integration for ergonomics and instructional clarity; and (3) evaluator experience.

The neurological examination followed several standards including essentials of neurological clinical assessments,^4,20^ the Part III of the Movement Disorders Society Unified Parkinson’s Disease Rating Scale (MDS-UPDRS),^21^ and video-based assessments developed or implemented in practice during the COVID-19 pandemic.^2,14^

### The examination

Using the standard iPad™ video app connected to a secure repository (Box™), the video exam technique, position, potential findings and duration are listed in Table 1. All sites were instructed to conduct the examinations at 3 standardized distances from the iPad: (1) approximately 2-3 feet for assessment of eye movements, facial expression, and speech evaluations, allowing for a focus on the face and neck; (2) most other physical movements were assessed at a distance of 8-10 feet which allowed for view of the entire participant; (3) standing up, posture, and gait assessments occurred over longer distances, walking up to 20 feet away from the camera (See Figure 1).

**Figure 1.**
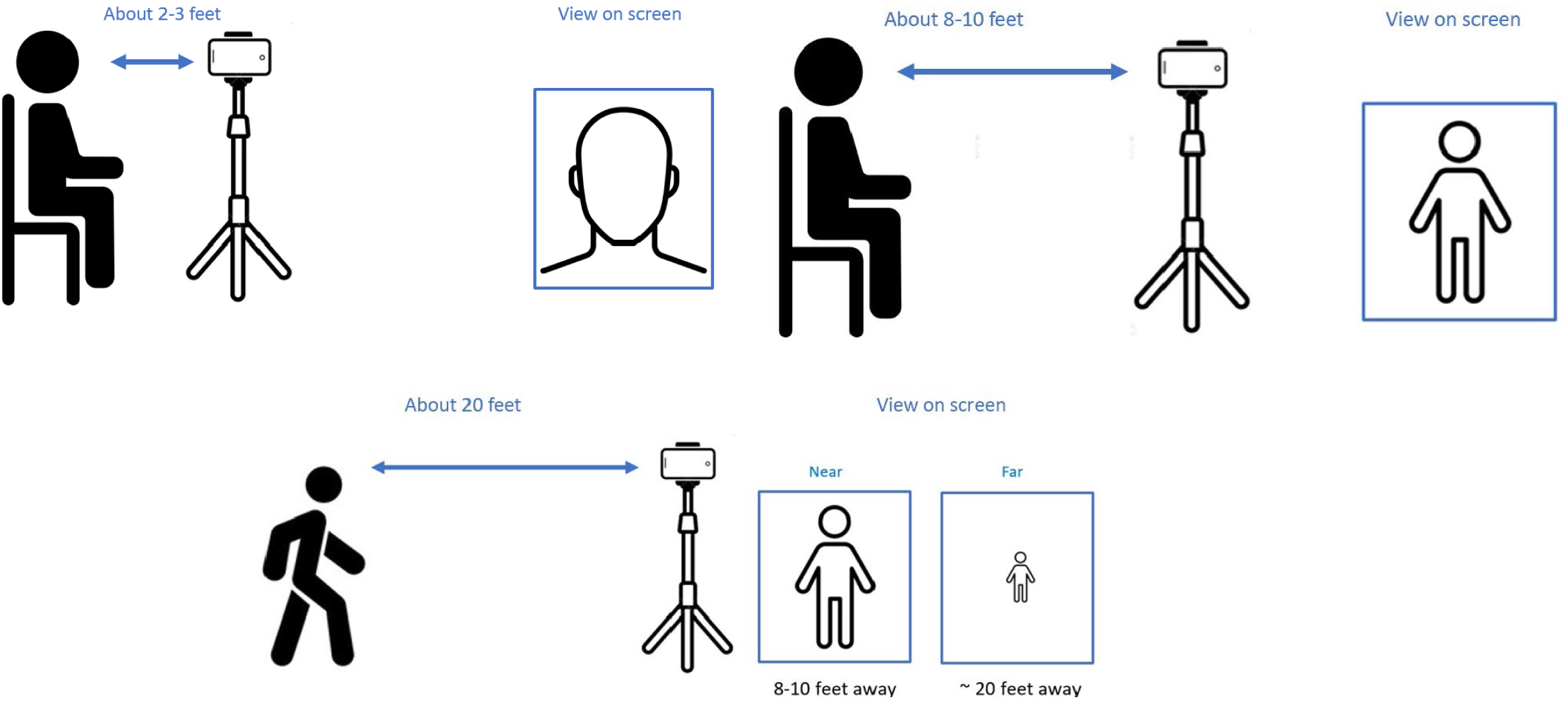
Camera positioning for VANE.

**Table 1.** Examination technique, position, and anticipated findings.

| Exam technique | Position relative to camera | Potential neurological findings | Duration spent in examination. |
| --- | --- | --- | --- |
| Extraocular movements in cardinal directions (cross pattern) | 2-3 feet | Monocular or binocular gaze restriction | 45 seconds |
| Rapid horizontal and vertical eye movements | 2-3 feet | Slow gaze initiation velocity | 25 seconds |
| Smile, eyebrow raise, eye closure | 2-3 feet | Facial weakness | 20 seconds |
| Articulation testing (labial sounds: “ma-ma”, “pa-pa”, lingual sounds: “la-la”, “ta-ta”, pharyngeal sounds: “ga-ga, ka-ka”) | 2-3 feet | Identification of dysarthria and potential lingual, labial or pharyngeal pattern | 10 seconds |
| Shoulder shrug, neck flexion, head turn | 2-3 feet | Sternocleidomastoid/related neck musculature weakness | 30 seconds |
| Cookie Theft interpretation | 2-3 feet | Spontaneous speech and language, visual neglect | 30-60 seconds |
| Pronator drift for 5s | 8-10 feet | Subtle corticospinal tract injury | 10 seconds |
| Finger to nose testing | 8-10 feet | Dysmetria or dyssynergia | 10 seconds |
| Arm raise, individual leg raise | 8-10 feet | gross motor strength (MRC ≥4/5) | 30 seconds |
| Rapid alternating movements (hand pronation/supination), 10 times | 8-10 feet | Rapidity/slowness of gross motor movements | 30 seconds |
| Resting hand position (10 count) | 8-10 feet | Resting tremor | 20 seconds |
| Rapid hand open/close (5 times each side) | 8-10 feet | Rapidity/slowness/arrests of gross motor movements | 20 seconds |
| Heel strike (6 inches off the floor, 5 times each side) | 8-10 feet | Rapidity/slowness/arrests of gross motor movements | 20 seconds |
| Finger taps, (10 times each side) | 8-10 feet | Rapidity/slowness/arrests of fine motor movements | 45 seconds |
| Praxis testing (slicing a loaf of bread, brushing teeth, salute, waving hello) | 8-10 feet | Assessment of complex motor tasks/praxis | 25 seconds |
| Modified get up and go | 10-20 feet | Ability to stand unassisted/assisted with hands | 15 seconds |
| Lateral posture (5 count) | 20 feet | Stooping of posture | 10 seconds |
| 20 foot walk trials (2 cycles away from and to camera) | 20 feet | Speed of walking, symmetry of limb movements, arm swing | 60 seconds |

### Data collection

Nearly all elements of the UDSv3 B8 form were adapted for inclusion in the DPPOS-4 form (“NB8”, Supplement). Rigidity and pull-testing were not included following the principle that PCs would not execute physical examination procedures requiring contact. Postural stability was inferred based on a modified get up and go test, a static postural position briefly held perpendicular to the camera, and other observations during the examination.

Additionally included in the assessment form were abnormal findings not typically associated with aging but known complications of microvascular ischemic neural injuries in diabetes mellitus including focal unilateral extraocular movement abnormality (an extraocular deficit in CN3, 4, or 6) and Bell’s palsy. Other common non-neurological examination findings were recorded including joint deformities (or gait abnormality due to orthopedic cause), difficulties standing or multiple attempts to stand up from sitting, and a general “other” option which enabled free-text description of findings not otherwise captured by this form. Evidence of complete hemifacial weakness or hemifacial synkinesis were considered as evidence of history of a Bell palsy.

### Training and Certification

To prepare site study staff, a 10-minute training video was developed to demonstrate the examination step-by-step. VANE could be conducted using written scripts read aloud by on-site staff (available in English and Spanish) or following a version of the staff instruction video shown to participants in real time as the video exam was recorded, allowing participants to follow the recorded neurologist’s instructions and mimic his movements.

Research staff directed all script-based VANEs and remained with participants for video-led VANEs. PCs were encouraged to repeat any segment of the examination for which the participants had trouble following standard instruction, or coach them to the desired action or position, demonstrating desired positions and movements in the process. This involved positioning of hands for observation of possible resting tremor, or when encouraging rapidly alternating movements, finger taps, or heel strikes to be as fast as possible for a count of ten (rather than the participant simply demonstrating they precisely could do an action 10 times well). PCs used available research space utilized for DPPOS-4 assessments for the VANE.

Site-specific and staff-specific performance review, feedback, and staff certification preceded central reading of video recordings by 3 physicians. Given the nature of the program, ongoing feedback was possible for each video if drift in examination quality occurred. An action plan was also developed for response to any clinical concerns identified during the assessment.

### Data Capture and Centralized Physician Review

Following the clinical examination, video recordings were uploaded to a HIPAA compliant Box repository. The completion of each exam was documented on the Visit Inventory Form (“P16” form; see Supplement) with the video embedded within the form which triggers an automated evaluation workflow managed by the Multimodal Integrated Data Acquisition System (MIDAS, CC-GWU). MIDAS synchronized the video files with corresponding NB8 clinical forms and managed physician assignments.

To ensure specialized oversight, examinations were randomly assigned among three physicians experienced in standardized examination techniques; participants requiring Spanish-language evaluation were assigned to two physicians fluent in Spanish. Continuous recording was encouraged to capture coincidental clinical signs in a single segment, but the platform supported the integration of up to three segments (Cranial Nerves, Motor Exam, and Gait/Balance). The MIDAS interface enabled non-linear navigation, allowing reviewers to move fluidly between sections to correlate findings across different exam components. Videos could be accelerated, slowed, moved forward or backward as required by the adjudicator. In cases of equivocal findings, the platform facilitated secure inter-reviewer communication for secondary review, with all final assessments adjudicated and recorded directly within MIDAS.

### Return of abnormal findings to participant

Upon identification of a concerning finding by the physician, the core manager and the CC-GWU were informed of the physician’s impression and advised action plan. Urgency in notifying the participants of the neurological abnormality was marked on the neurological exam form filled by the physician interpreting the VANE. Site research staff were then notified to review the findings and reconcile with any available research records or other clinical records which can be provided. If a finding was known to the participant but not previously recorded in DPPOS, this was documented without further action needed. If the findings identified by the physician were new and requiring further attention, the site PI or clinician reviewed the concerns to determine appropriate steps. Anticipated actions included a phone call to the participant by the site PC, encouraging follow-up/review with primary care physician. Support by the VANE reviewing physician and support was available to the site.

### Experience from Implementation

Over two years of implementation (11/2022 to 10/2024), 1286 VANEs were completed on participants completing in-person assessment, leading to 1284 examination reviews (Figure 2). Most VANEs (1204, 93%) were completed using a script at all 25 DPPOS-4 sites. Additionally, 78 (6%) of examinations were completed following the video recorded prompts at 10 sites. Two sites completed more than half of their evaluations with the video recorded prompts. Most examinations (1237, 96%) were delivered as planned, 41 (3%) had minor errors but were usable and 6 (0.4%) had major deviations in examination and an examination impression could not be determined. Only one research staff member needed more than one feedback session. An additional two examinations could not be reviewed because recorded videos were inaccessible due to technical errors. Lower quality examinations were from early implementation (before feedback was given). No site had more than 2 unusable visits. Each site on average completed 51.4 examinations (range 14-92).

**Figure 2.**
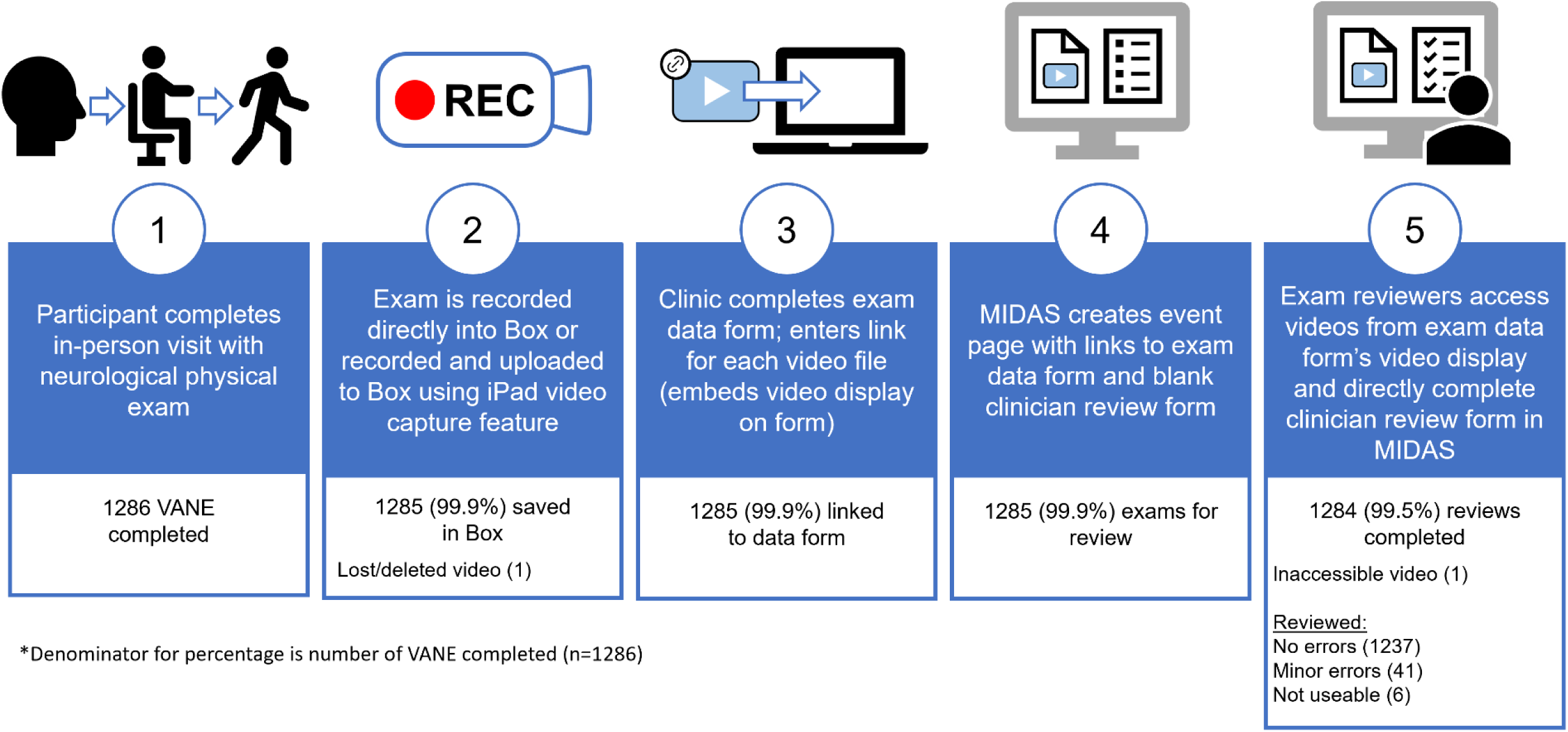
DPPOS VANE Procedure, Review, and Assessments.

Videos were on average 10 minutes long with variability related to transitions between examination sections and spaces (e.g. transition from a room to a corridor for the gait exam). At 10 minutes per video, and 1284 videos reviewed, approximately 214 hours of videos were recorded, on average 8.6 hours of video per study site. Dividing the work by the 3 physician reviewers, each person reviewed approximately 71 hours of videos.

## Discussion

Engaging 55 research staff, and 3 physician-reviewers across 25 sites, this study is the first to demonstrate VANE as an efficient neurological examination model suited for studies and settings in which harmonized, reliable and comprehensive neurologic examinations are traditionally considered infeasible, using commonly available technology and study personnel.

Relative to a conventional model whereby each site has a local expert, the VANE approach eliminated the need for 22 additional clinician experts. This represents a dramatic reduction in required personnel to complete this work along with a substantial savings in cost and time spent in training. For comparison, a traditional model of 25 sites would have required 22 additional clinician experts, who would likely save about 5 minutes for each exam,^11^ but that time does not take into account many other factors including time spent coincident to a visit, and complicated schedules of busy clinicians. Moreover, the approach taken assures a consistency in the amount of video reviews completed by the 3 evaluating clinicians, which is important given a wide range in each site’s cohort size, translating to range from approximately 2.2 to 15.3 of hours of video per site. Finally, the asynchronous manner of neurological exam review provides greater flexibility for study implementation and schedules which may not align with the site, particularly when spread across multiple time zones as is the case in DPPOS.

VANE presents a novel paradigm for multisite epidemiological studies considering neurological examinations, engaging a small team of clinical and research staff and addressing barriers to evaluation including geography and personnel. VANE could be implemented beyond research settings including use as a platform for clinical evaluation at long-term care facilities, drug monitoring, supplementation with telemedicine, and serve as resource for assessing neurological status in other settings when healthcare providers are not on site. VANE recordings also provide capabilities for automated assessments including AI-trained models to track longitudinal changes in speech, articulation, and motor exam features, which may be predictive of cognitive changes in AD/ADRD^22^ and motor changes in parkinsonism.^23^

VANE has several limitations. First, tone testing cannot be assessed and this is a recognized limitation of remote evaluations. Visual fields were not formally conducted, but are recognized as being poorly sensitive and likely uninformative when conducted in a screening manner.^24^ While the *Cookie Theft* picture enables a participant to review picture gestalt, it is unlikely to identify a pure hemianopsia, but does provide an observable example of language examination beyond brief responses to the examiners.

The development of VANE has overall been successful in DPPOS and generally met the goals of the program. We believe that VANE can be implemented in other multisite studies in a cost-efficient manner. A second wave of neurological evaluations are underway. Beyond the methods described above, a full report of neurological examinations among participants will follow, aiming to provide insights into the prevalence and progression of neurological findings in the DPPOS-4 cohort of older adults with prediabetes or diabetes.

## Supporting information

DPPOS Investigators Appendix

NB8 Form

P16 Form

## Acknowledgement

The authors wish to acknowledge the exceptional commitment of DPPOS participants and study staff to put this new approach into practice.

## Statement of Ethics

### Study approval statement

This study protocol was reviewed and approved by the Advarra single IRB (protocol Pro00047977)

Consent to participate statement: Written informed consent was obtained from participants (or their parent/legal guardian/next of kin) to participate in the study.

### Funding Sources

DPPOS was funded by grant U19AG078558 from the National Institutes of Health.

DPP/DPPOS NIH program officers have participated in DPP/DPPOS Steering and Executive Committees meetings which designed the study, and the PPS that oversaw reporting of DPP. No NIH member had a direct role in data collection, data analysis, writing, or reporting of this manuscript

This project was supported by the National Institute of Diabetes and Digestive and Kidney Diseases (NIDDK) of the National Institutes of Health (NIH) under award numbers U01 DK048489, U01 DK048339, U01 DK048377, U01 DK048349, U01 DK048381, U01 DK048468, U01 DK048434, U01 DK048485, U01 DK048375, U01 DK048514, U01 DK048437, U01 DK048413, U01 DK048411, U01 DK048406, U01 DK048380, U01 DK048397, U01 DK048412, U01 DK048404, U01 DK048387, U01 DK048407, U01 DK048443, and U01 DK048400, and in part by the National Institute on Aging of the NIH under award number 5 U19 AG078558 by providing funding during DPP and DPPOS to the clinical centers and the Coordinating Center for the design and conduct of the study, and collection, management, analysis, and interpretation of the data. Funding was also provided by the National Institute of Child Health and Human Development, the National Eye Institute, the National Heart Lung and Blood Institute, the National Cancer Institute, the Office of Research on Women’s Health, the National Institute on Minority Health and Health Disparities, the Centers for Disease Control and Prevention, and the American Diabetes Association. The content is solely the responsibility of the authors and does not necessarily represent the official views of the National

Institutes of Health. The Southwestern American Indian Centers were supported directly by the NIDDK, including its Intramural Research Program, and the Indian Health Service. The General Clinical Research Center Program, National Center for Research Resources, and the Department of Veterans Affairs supported data collection at many of the clinical centers. Merck KGaA provided medication for DPPOS. DPP/DPPOS have also received donated materials, equipment, or medicines for concomitant conditions from Bristol-Myers Squibb, Parke-Davis, and LifeScan Inc., Health O Meter, Hoechst Marion Roussel, Inc., Merck-Medco Managed Care, Inc., Merck and Co., Nike Sports Marketing, Slim Fast Foods Co., and Quaker Oats Co. McKesson BioServices Corp., Matthews Media Group, Inc., and the Henry M. Jackson Foundation provided support services under subcontract with the Coordinating Center. The sponsor of this study was represented on the Steering Committee and played a part in study design, how the study was done, and publication. All authors in the writing group had access to all data. The opinions expressed are those of the study group and do not necessarily reflect the views of the funding agencies. A complete list of Centers, investigators, and staff can be found in the Appendix.

### Author Contributions

Conceptualization: JAL, JMN

Data curation: MT, AB, LD

Formal analysis: LD, MT

Funding acquisition: JAL, JMN

Investigation: All

Methodology: JAL, JMN, NN

Resources: JL, MT

Software: MT, LD, AB

Supervision: JAL, JMN

Validation: MT, LD, AB

Writing – original draft: JMN, JAL

Writing – review & editing: JMN, JAL, NK, MT, OC, GT, VS

### Data Availability Statement

In accordance with the NIH Public Access Policy, we continue to provide all manuscripts to PubMed Central including this manuscript DPP/DPPOS has provided the protocols and lifestyle and medication intervention manuals to the public through its public website (https://www.dppos.org). The DPPOS abides by the NIDDK data sharing policy and implementation guidance as required by the NIH/NIDDK (https://repository.niddk.nih.gov/studies/dppos/)

