## Supplementary material for "Implementation of a standardized Video-based Asynchronous Neurological Examination (VANE) in a multi-center observational study of Alzheimer’s disease (AD) and AD related dementias": NB8 Form

Diabetes Prevention Program Outcomes Study

### P16 NEUROLOGICAL PHYSICAL EXAM ASSESSMENT

**This form is used by clinic staff to document the Neurological Physical Exam assessment completed at each Main Visit. This form should be completed for all participants, even if the Neurological Physical Exam assessment was not attempted. Follow procedures in the Neurological Physical Exam manual for the administration, set-up, capture, and transmission for the assessment.**

**If the Neurological Physical Exam assessment was not attempted, complete Section A of the form.**

A. Visit Information

1. Outcome visit

2. Date of visit — month / day / year

3. Was the Neurological Physical Exam assessment attempted? Yes [1] No [2]

**If NO,**

a. What was the main reason why the Neurological Physical Exam assessment was not attempted?

***CHECK ONLY ONE***

Participant refused to complete [1]
Participant did not have enough time to complete [2]
Cognitive impairment prevents completion [3]
Physical impairment prevents completion [4]
Other [5]

1. **If OTHER**, specify:

**If YES,**

b. Start time of assessment __ __ : __ __
24 hour clock

c. The Neurological Physical Exam was administered:

***CHECK ONE ONLY***

Live, with provided script [1]
Using video prompts [2]

Identification code of person administering assessment | Form entered in computer?

B. Neurological Physical Exam Assessment Completion

1. The Neurological Physical Exam assessment was: ***CHECK ONLY ONE***

Completed without problem [1]

Completed **with** problem [2]

**Not completed** [3]

**If 'Completed with problem' or 'Not completed,'**

a. What was the main reason? ***CHECK ONLY ONE***

Participant refused to complete [1]

Participant did not have enough time to complete [2]

Cognitive impairment prevents completion [3]

Physical impairment prevents completion [4]

Technical issues prevents completion [5]

Other [6]

1. **If OTHER,** specify:

**If the Neurological Physical Exam assessment was attempted but NOT COMPLETED, SKIP to section D. If the Neurological Physical Exam assessment was COMPLETED WITHOUT PROBLEM or COMPLETED WITH PROBLEM, CONTINUE to section C.**

C. Transmission Information

**The Neurological Physical Exam assessment for asynchronous review should be recorded as a single video file. The file should be recorded directly to the participant's folder in BOX or uploaded to the participant's folder in BOX. The file name and the shared link for the file will be entered in section C. If multiple video files were recorded, include all relevant files/links in question C2 (up to 5 video files).**

1. Date video files recorded/uploaded to clinic's Box folder — month / day / year

2. File upload information from BOX

| File | File name of video from BOX | URL of video file from BOX |
|---|---|---|
| a. Video file 1 | | [Entered in MIDAS only] |
| b. Video file 2 | | [Entered in MIDAS only] |
| c. Video file 3 | | [Entered in MIDAS only] |

| | | |
|---|---|---|
| d. Video file 4 | | [Entered in MIDAS only] |
| e. Video file 5 | | [Entered in MIDAS only] |

3. Clinic confirms correct participant videos are displayed in MIDAS after saving the form? Yes [1] No [2]

D. Comments

1. Indicate any comments or problems regarding the administration, completion, or data transmission/upload of this assessment:

______________________________

______________________________

______________________________

______________________________

______________________________

______________________________
