## Supplementary material for "Implementation of a standardized Video-based Asynchronous Neurological Examination (VANE) in a multi-center observational study of Alzheimer’s disease (AD) and AD related dementias": P16 Form

SAMPLE FORM

### NB8 [V1]: NACC Neurological Examination Findings

Smart Skip: ☑ Cancel

**Clinic:** **Participant ID:** **Nickname:** **Sex:**

**Neurological physical exam review number** NB8

#### A. VISIT INFORMATION

1. Outcome visit for assessment
2. Date of assessment
3. P16 Handle
4. Date of review
5. Initials of reviewer
6. The Neurological Physical Exam review was completed:
   - ☐ 1=With PC using provided script
   - ☐ 2=With PC using video prompts
   - ☐ 3=With neurologist (Clinics 13 or 21 for validation)
7. Were any physical or hearing aids used during the assessment? ☐ Yes ☐ No

**If YES,**

**CHECK ALL THAT APPLY**

a. Walking aid or gait assistance device ☐ 1=Yes

b. Hearing aid ☐ 1=Yes

c. Other ☐ 1=Yes

**If OTHER,**

1. Specify:

#### B. NEUROLOGICAL EXAMINATION FINDINGS EVALUATION

**INSTRUCTIONS: This form must be completed by a clinician with experience in assessing the neurological signs listed below and in attributing the observed findings to a particular syndrome. Please use your best clinical judgment in assigning the syndrome. For additional clarification and examples, see UDS Coding Guidebook for Initial Visit Packet, Form B8.**

1. Were there abnormal neurological exam findings?
   - ☐ 0=No abnormal findings
   - ☐ 1=Yes abnormal findings were consistent with syndromes listed in Questions 2-8
   - ☐ 2=Yes abnormal findings were consistent with age-associated changes or irrelevant to dementing disorders (e.g., Bell's palsy)

**If Question 1 is NO ABNORMAL FINDINGS (option 0), SKIP to Section C. If Question 1 is YES, ABNORMAL FINDINGS CONSISTENT WITH AGE-ASSOCIATED CHANGES OR IRRELEVANT TO DEMENTING DISORDERS (option 2), SKIP to Question 8. Otherwise, CONTINUE.**

**INSTRUCTIONS FOR QUESTIONS 2 - 8**

**Please complete the appropriate sections below, using your best clinical judgment in selecting findings that indicate the likely syndrome(s) that is/are present.**

**CHECK ALL OF THE GROUPS OF FINDINGS / SYNDROMES THAT WERE PRESENT :**

2. Parkinsonian signs ☐ 0=No ☐ 1=Yes

**If YES,**

| | LEFT | | RIGHT | |
|---|---|---|---|---|
| 2a. Resting tremor - arm | ☐ 1=Yes | ☐ 8=Not assessed | ☐ 1=Yes | ☐ 8=Not assessed |
| 2b. Slowing of fine motor movements | ☐ 1=Yes | ☐ 8=Not assessed | ☐ 1=Yes | ☐ 8=Not assessed |
| 2c. Rigidity - arm | ☐ 1=Yes | ☐ 8=Not assessed | ☐ 1=Yes | ☐ 8=Not assessed |

| | | |
|---|---|---|
| 2d. Bradykinesia | ☐ 1=Yes | ☐ 8=Not assessed |
| 2e. Parkinsonian gait disorder | ☐ 1=Yes | ☐ 8=Not assessed |
| 2f. Postural instability | ☐ 1=Yes | ☐ 8=Not assessed |

3. Neurological signs considered by examiner to be most likely consistent with cerebrovascular disease ☐ 0=No ☐ 1=Yes

**If YES,**

| | | |
|---|---|---|
| 3a. Cortical cognitive deficit (e.g., aphasia, apraxia, neglect) | ☐ 1=Yes | ☐ 8=Not assessed |
| 3b. Focal or other neurological findings consistent with SIVD (subcortical ischemic vascular dementia) | ☐ 1=Yes | ☐ 8=Not assessed |

| | LEFT | | RIGHT | |
|---|---|---|---|---|
| 3c. Motor (may include weakness of combinations of face, arm, and leg; reflex changes; etc.) | ☐ 1=Yes | ☐ 8=Not assessed | ☐ 1=Yes | ☐ 8=Not assessed |
| 3d. Cortical visual field loss | ☐ 1=Yes | ☐ 8=Not assessed | ☐ 1=Yes | ☐ 8=Not assessed |
| 3e. Somatosensory loss | ☐ 1=Yes | ☐ 8=Not assessed | ☐ 1=Yes | ☐ 8=Not assessed |

4. Higher cortical visual problem suggesting posterior cortical atrophy (e.g., prosopagnosia, simultagnosia, Balint's syndrome) or apraxia of gaze ☐ 0=No ☐ 1=Yes

5. Findings suggestive of progressive supranuclear palsy (PSP), corticobasal syndrome, or other related disorders ☐ 0=No ☐ 1=Yes

**If YES,**

| | | |
|---|---|---|
| 5a. Eye movement changes consistent with PSP | ☐ 1=Yes | ☐ 8=Not assessed |
| 5b. Dysarthria consistent with PSP | ☐ 1=Yes | ☐ 8=Not assessed |
| 5c. Axial rigidity consistent with PSP | ☐ 1=Yes | ☐ 8=Not assessed |
| 5d. Gait disorder consistent with PSP | ☐ 1=Yes | ☐ 8=Not assessed |
| 5e. Apraxia of speech | ☐ 1=Yes | ☐ 8=Not assessed |

| | LEFT | | RIGHT | |
|---|---|---|---|---|
| 5f. Apraxia consistent with CBS | ☐ 1=Yes | ☐ 8=Not assessed | ☐ 1=Yes | ☐ 8=Not assessed |
| 5g. Cortical sensory deficits consistent with CBS | ☐ 1=Yes | ☐ 8=Not assessed | ☐ 1=Yes | ☐ 8=Not assessed |
| 5h. Ataxia consistent with CBS | ☐ 1=Yes | ☐ 8=Not assessed | ☐ 1=Yes | ☐ 8=Not assessed |
| 5i. Alien limb consistent with CBS | ☐ 1=Yes | ☐ 8=Not assessed | ☐ 1=Yes | ☐ 8=Not assessed |
| 5j. Dystonia consistent with CBS, PSP, or related disorder | ☐ 1=Yes | ☐ 8=Not assessed | ☐ 1=Yes | ☐ 8=Not assessed |
| 5k. Myoclonus consistent with CBS | ☐ 1=Yes | ☐ 8=Not assessed | ☐ 1=Yes | ☐ 8=Not assessed |

6. Findings suggesting ALS (e.g., muscle wasting, fasciculations, upper motor neuron and/or lower motor neuron signs)
   - ☐ 0=No
   - ☐ 1=Yes

7. Normal-pressure hydrocephalus: gait apraxia
   - ☐ 0=No
   - ☐ 1=Yes

8. Other findings (e.g., cerebellar ataxia, chorea, myoclonus)
   (NOTE: For this question, do not specify symptoms that have already been checked above)
   - ☐ 0=No
   - ☐ 1=Yes

**If YES,**

**CHECK ALL THAT APPLY**

a. Focal unilateral extra occular movement abnormality (Deficit in CN3, 4, or 6) ☐ 1=Yes
b. Bell's Palsy ☐ 1=Yes
c. Joint deformities (or gait abnormality due to orthopaedic cause) ☐ 1=Yes
d. Difficulties standing or multiple attempts to stand up from sitting ☐ 1=Yes
e. Other ☐ 1=Yes

**If OTHER,**
1. Specify:

##### C. FINAL IMPRESSIONS

1. Indicate any problems that may have affected the participant's completion of the neurological physical exam

**CHECK ALL THAT APPLY**

a. Hearing problem ☐ 1=Yes
b. Comprehension problem ☐ 1=Yes
c. Physical problem ☐ 1=Yes
d. Technical problem ☐ 1=Yes
e. Other ☐ 1=Yes

**If OTHER,**
1. Specify:

2. Record any final comments or other general impressions of the assessment.

3. Review the administration of the examination/assessment. These results will be used as part of the Neurological Physical Exam quality control review.
   - ☐ 1=Full neurological examination, with few or no deviations from the script, and there was no impact on interpretation of the examination.
   - ☐ 2=Neurological examination had a substantial number of deviations, but was still of sufficient quality such that a determination of normal examination or a neurological diagnosis could be conferred with sufficient examiner confidence.
   - ☐ 3=Neurological examination not interpretable.

**If neurological examination was NOT INTERPRETABLE, specify why:**

**CHECK ALL THAT APPLY**

a. Technical problems with recording ☐ 1=Yes
b. Inadequate examination technique ☐ 1=Yes

c. Incomplete video recording ☐ 1=Yes

d. Other ☐ 1=Yes

**If OTHER,**

1. Specify:

4. Were there any concerns regarding the participant's assessment that necessitate or require clinic follow-up with the participant or their PCP? ☐ Yes ☐ No

**If YES,**

a. What is the urgency of the concern?

☐ 1=Needs attention (not urgent)

☐ 2=Needs urgent attention

b. Describe the concern:
